# Gastrointestinal parasites of zoonotic importance detected in bats in the conservation area of Semuliki National Park, western Uganda

**DOI:** 10.1101/2025.04.10.25325607

**Authors:** James Robert Ochieng, Charles Drago Kato, John Joseph M. Kisakye

## Abstract

Bat guano may contain zoonotic parasites that contaminate the environment and/or serve as a potential source of infection to humans and animals. Repeated bat-human exposure could be a risk factor for zoonosis. To date, knowledge on the status of bat gastrointestinal parasites (GIPs) in Uganda is limited. We conducted a cross-sectional study to investigate the prevalence of bat GIP species in communities contiguous to Semuliki National Park (SNP) Bundibugyo district in western Uganda. We purposively collected faecal samples of micro- (n=242) and mega-bats (n=242) from bat roosts in communities contiguous to SNP during the rainy months of October to December 2023 and the dry months of January to March 2024. Standard faecal floatation and sedimentation techniques were used for laboratory examination. Microscopic examination revealed that 224 (46%) samples tested positive for more than one parasite species. Thirteen GIPs, including protozoa (n=3), trematode (n=1), cestode (n=1), and nematodes (n=8) were detected. The most prevalent parasites were *Entamoeba coli* (57%), Hookworm (33%), *Strongyloides* sp. (33%), and *E*. *histolytica* (32%), and the least prevalent were the two Unidentified nematodes (1%). 77% (n=10) of the detected GIPs are responsible for zoonosis and are of significant public health importance. Statistically, there was a significant difference (*P* < 0.05) in the overall parasite prevalence across the four studied bat groups. Also, parasite prevalence was significantly higher in microbats compared to megabats (*P* < 0.05) and in Burondo sub-county compared to Ntandi town council (*P*< 0.05). The detected zoonotic parasites pose a potential source of zoonosis in communities contiguous to the conservation area of Semuliki National Park, Uganda. This calls for awareness creation on the risks of bat mediated zoonotic parasitosis and the use of good sanitary practices to prevent chances of zoonotic parasite spillover from bats to humans.

## Background

Gastrointestinal parasitic infections in wildlife can be influenced by abiotic and biotic factors and parasites’ ecology [1, 2]. GIPs play a major role in ecosystems, affecting hosts’ ecology and evolution of interspecific interactions, population growth and fitness, increasing vulnerability to diseases, and/or fatality if not treated [3–6]. Globally, GIPs have been recognized as causing significant morbidity and mortality in wild fauna, including bats [3, 7, 8], and are therefore essential parasites to consider in wildlife conservation [5]. The current climate change, along with other drivers like an increase in human population, changes in land use, road construction projects, forest penetration and destruction of parasite reservoirs’ natural habitats, natural calamities, and illegal hunting, threatens bat populations [7]. These factors increase the risks of human-bat interaction, yet bats play a significant role in pathogen transmission [6, 8].

Bats (order Chiroptera), are the only active flying true placental mammals of the animal kingdom and are the second largest order of mammals after rodents (order Rodentia) with cosmopolitan distribution [14, 15]. Traditionally, bats are classified into two major groups: suborder megachiroptera (megabats), the fruit eating bats (fruit bats), and suborder microchiroptera (microbats), the insectivorous bats [16, 17].

The megabats are much larger herbivores, and they consume plant fruits, flowers, leaves, nectar, and pollens, and are commonly seen in fruiting trees where they roost in tightly packed clusters [14–17]. In contrast, the microbats are mostly insectivorous, though a few of these species may also feed on blood, fruits, nectars, pollens, and vertebrates [14, 15]. Microbats are more common and numerous than megabats and usually hang in caves, on roofs, and in tree hollows during the day [15].

Globally, more than 1,400 bat species have been reported [8, 18], although many are currently threatened, and over 289 species are categorised as endangered, vulnerable, or near threatened by the International Union for Conservation of Nature (IUCN) Red List [8]. Studies by [17] and [18] documented 90 bat species: 13 mega- and 77 micro-bats in Uganda, with over 10 species suspected to be in the conservation areas of Semuliki Bundibugyo district western Uganda. In Uganda, bats are widespread throughout the country and are particularly abundant in the conservation areas, suburban and urban areas, including in domestic settings, human buildings, hospitals, church and school premises [19]. Other bat roosts are known to inhabit different environmental settings, including caves, disused mines, rock crevices, tree hollows and holes, and termite nests [19, 20]. However, the bats’ ecology varies and is peculiar with their feeding behaviour, which also determines the risks of bat-parasite and bat-human interactions [8].

In the aspect of public health, over 70% of new, emerging, and reemerging infectious diseases are of animal origin, and research has shown that bats play a central role in the ecosystem by serving as carriers, reservoirs, and/ or transmitters of pathogens of public health importance globally [6]. The bat-transmitted pathogens include fungi, bacteria, viruses, and parasites, which can affect humans and/ or animals [6, 8, 21]. Bats may transfer these pathogens over long distances as they move from sylvatic to domestic settings and vice versa while seeking food and other basics and as they share shelter with humans and other animals [6, 8].

Currently, Uganda has documented a total of several bat-mediated pathogens, including numerous rabies, Marburg viral hemorrhagic fever [22], and eight Ebola outbreaks involving the districts of Gulu (2000), Bundibugyo (2007), Luwero (2011 & 2012), Kibaale (2012), Luwero (2012), Mubende and Kasanda (2022), and Kampala (2025) since 2000 [19, 23, 24]. The recent Marburg outbreak in the Kween district, Eastern Uganda, was traced to rock salt mining in a bat cave [19], and the Egyptian fruit bat, *Rousettus aegyptiacus* was identified as a Marburg virus reservoir [25]. However, these zoonotic bat-associated pathogenic viruses still need more attention for human and animal health in Uganda and beyond and are also critical for wildlife conservation.

Bat parasitosis studies in East Africa, including Kenya [15], other African countries [26, 27], and beyond Africa, documented several GIPs, including those responsible for zoonosis. Studies by [28] reported 59 species of helminths: 28 nematodes, 23 trematodes, 6 cestodes, and 2 acanthocephalans in Brazil. [15] reported *Ascarid* spp., *Capillarid* sp., *Cryptosporidium* sp., *Eimeria* spp., *Entamoeba* sp., Giardia sp., *Hymenolepis* spp., *Isospora* sp., *Oxyurid* sp., *Strongyle*, and *Strongyloides* sp. in insectivorous bats and *Eimeria* sp., *Entamoeba* sp., and *Hymenolepis* sp. in frugivorous bats in Kenya. [27] detected 5 nematodes, 2 trematodes, and 2 cestodes in microbats in southeast Nigeria. [26] described several nematodes, trematodes, and cestodes in micro- and mega-bats in Egypt. [29] found *Cryptosporidium* spp. in bats from the USA and the Czech Republic. [30] and [31] observed *Hymenolepis* spp. in bats in China and Japan, respectively. [32] discovered 2 *Eimeria* spp. in microbats of the family Vespertilionidae: eastern red bat (*Lasiurus* borealis) in North Carolina, while [33] recognized 6 *Eimeria* spp. in Vespertilionid bats in North America. The variations in bat GIP diversity and/or prevalence may be due to differences in the status of the sampled bats, geographical location, seasonal variations, and the diagnostic techniques used [6, 8]. Other related studies documented bat ectoparasites, including ticks, fleas, and bed bugs [8, 37–39].

Bat parasitic infections may range from minimal to advanced effects and can significantly affect their fitness, depressing their metabolism, as has been reported elsewhere [7, 8]. These parasitic infections suppress bats’ physiological and immunological responses, leading to reduced movement/flight capability, breeding success, and increased inactivity, increasing their vulnerability to predation and other diseases, resulting in death in most cases [6–8, 36]. This is critical for wildlife conservation and can lead to biodiversity loss in addition to public health risks.

Currently, several bat roosts consisting of over 300 bats have been reported in human settings: homes, schools, hospitals, churches, trees (like cocoa, mangoes, and avocadoes), disused mines, rock crevices, and termite nests in Burondo sub-county and Ntandi town council Bundibugyo district, Uganda by the Uganda Wildlife Authority, STOP Spillover team [19] and community members. This may worsen due to climate change, environmental shifts, among other factors. The bats pose a nuisance and health risks to humans. Surprisingly, up-to-date, little is known about the bat GIPs inhabiting the conservation area of SNP. Understanding the status of bat GIPs in communities contiguous to SNP is crucial for implementing effective control and prevention strategies for parasite zoonotic spillover from bats to humans. This study aimed to broaden knowledge on the current status of bat GIPs in bat hotspots in human communities contiguous to SNP to prevent chances of zoonotic parasite spillover from bats to humans. The objectives were to quantify the diversity and prevalence of bat GIPs and to assess the risks of bat mediated zoonotic parasitosis. Given the present limitations to national surveillance of wildlife-mediated parasitoses in Uganda, including scarce resources, focusing on forecasting spillover dynamics of bat-mediated zoonoses using a simple approach “bat guano diagnosis” could be a wise alternative. This study will contribute more knowledge on the bat GIP species diversity, and the risks of zoonotic parasitoses in communities contiguous to SNP Bundibugyo, Uganda. This will increase awareness of the risks of bat-human interaction in human communities to save lives and strengthen biodiversity conservation.

## Materials and methods

### Study area

The study was carried out in human communities in Burondo sub-county and Ntandi town council contiguous to SNP in Bundibugyo district, western Uganda. The study area, being contiguous to the park, possesses a high level of human-wildlife-forest ecosystem interactions. Bundibugyo landscapes have plenty of rock shelters and caves that are habitats for wildlife, including bats [19]. Bundibugyo receives an average rainfall of 1,250 mm, with rainfall peaks from March to May and from September to December, characterized by excessive flooding.

SNP covers an area of 220 km² and lies between 0“44’-0^0^53’N and 29“57’-30^0^11’E with an altitudinal range of 670-760 m above sea level and 18 to 30 °C (64 to 86 °F) temperature range, with relatively small daily variations. Semuliki Forest was made a National Park in October 1993, making it one of Uganda’s newest National Parks, and is the only remaining primary is contiguous with undisturbed forests of the Congo basin including Virunga National Park in the Democratic Republic of Congo to the west, and is known to be rich in biodiversity, possessing many butterfly species and more than 441 bird species, with 31 species including capuchin babbler, piping hornbill, blue-headed crested flycatcher, red-bellied malimbe and orange weaver among others, known to occur nowhere else in East Africa [39] and over 10 bat species. Due to the current increase in the human population in Uganda, SNP, among other protected areas, remains under threat by agriculturalists and wild hunters/poachers, despite the conservation efforts by the Uganda Wildlife Authority (UWA) and stakeholders. Preliminary investigation by the UWA, STOP Spillover team [19] and communities have identified Burondo sub-county and Ntandi town council as hotspot places with a significant population of bats. Furthermore, bat poachers have been reported by UWA in the communities of Burondo sub-county and Ntandi town council in contiguity to SNP. The bat poachers go to the park to collect firewood, poles, palm oil, and grass for domestic use.

**Figure 1:**
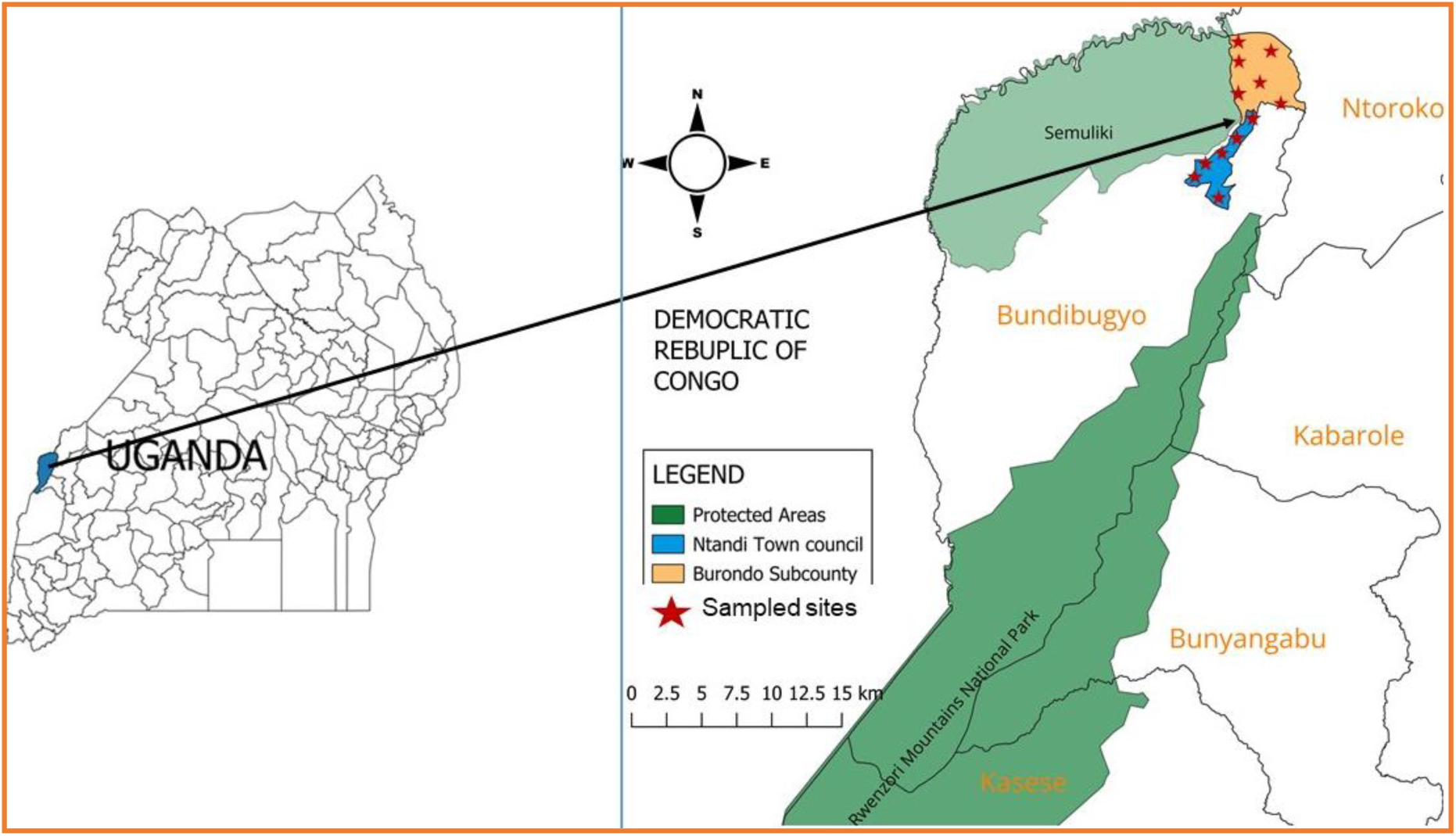
Map of the study area showing the location of Ntandi town council and Burondo sub-county in Bundibugyo district western Uganda. The Red stars show the sampled sites within the study area.

**Figure 2:**
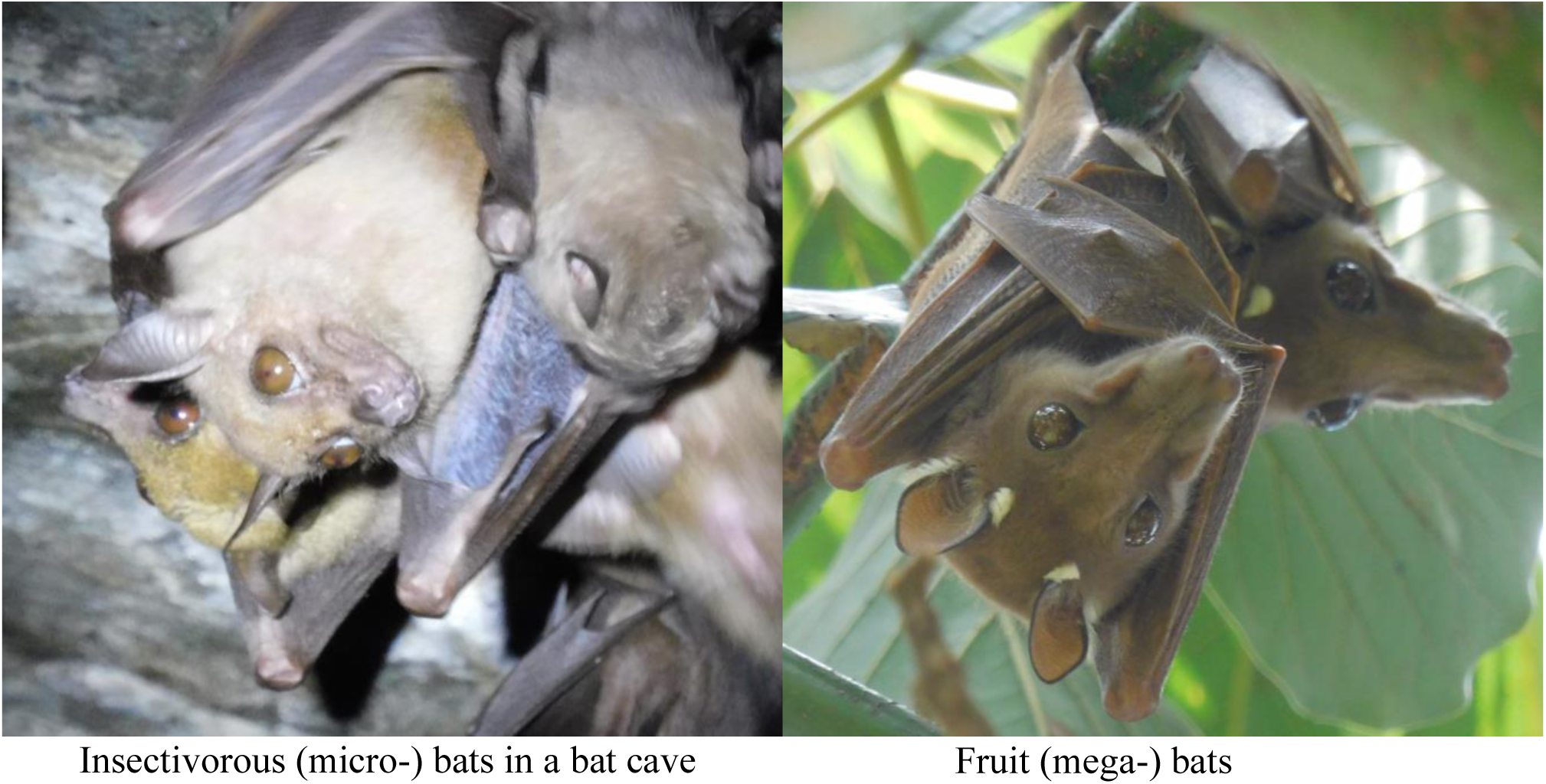
Photograph of a colony of micro-(insectivorous) bats in a roost resting in a bat cave (left) and two megabats (Egyptian fruit bats, *Rousettus aegyptiacus*) resting on an avocado tree in Burondo sub-county, Bundibugyo. *Original photos by James Robert Ochieng*.

### Study design and field faecal sample collection

We applied a cross-sectional study design. Batches of bat faecal samples were purposively collected from bat roosts in communities in Burondo sub-county and Ntandi town council in contiguity to SNP Bundibgyo district western Uganda. Samples were collected during the wet months of October to December 2023 and the dry months of January to March 2024. The sample size was determined using Thrusfield’s standard formula [40]. Data collection involved both bat faecal sample collection and the use of a questionnaire.

Briefly, during the bat faecal sample collection, at least a total of 40 clean white polyester sheets of cloth were overlaid on the floor of five bat roosts per sub-county or town council every morning. With the help of forceps, bat faecal samples that fell on the white polyester sheets were collected daily at midday and evening. Quality control, such as observing the presence of other mammals inside the bat roosts, was put into consideration. All the collected samples were placed in clean 10 ml sterile vials where it was thoroughly mixed with 10% formalin for preservation at room temperature until laboratory analysis at the Zoology Department Makerere University Kampala Uganda. Socio-demographic data and detailed information on the season, GPS coordinates, roost abundance, human economic activities, lifestyle hygiene, and sanitation in the study sites were all captured.

### Parasite identification

All the collected bat faecal samples were subjected to floatation and sedimentation standard methods [41] before microscopic observations. Briefly, for floatation, portions of faecal samples were suspended in a saturated solution of sodium nitrate (NaNO_3_) to separate protozoan cysts and helminth eggs from faecal samples. Four millilitres of the saturated NaNO_3_ solution were added to 1 ml of formalised faecal sample in a beaker and stirred with an applicator before being filtered through a gauze mesh into a second beaker. The filtrate was poured into a two millilitre vial and filled to the brim, forming a convex meniscus. A cover slip was gently placed on top of the meniscus, avoiding any air bubbles, and left undisturbed for at least 5 minutes. The cover slip was then carefully lifted upwards by a straight pull and gently placed on its face downwards on a labeled glass slide stained with Lugol’s iodine. The slide was then examined under the microscope at 10X and 40X objective lenses for parasite identification.

Sedimentation involved adding two millilitres of formalised faecal sample to five millilitres of formol-ether, mixed and filtered through a 350 μm gauze mesh. The filtrate was poured into a 15 ml centrifuge tube followed by 3 ml of diethyl ether to make 10 ml, sealed and vortexed as prescribed by [41]. The mixture was then centrifuged at 1500 rpm for 5 minutes to form 4 layers. The supernatant (top three layers) was discarded, leaving the sediment. About 10 µl of sediment was pipetted onto a glass slide, a drop of Lugol’s iodine stain was added and covered with a coverslip, and examined under the microscope as at floatation. The GIP eggs, cysts, and trophozoites were identified using established structural and morphological principles [21, 37].

### Statistical analysis

All the statistical tests were performed using the IBM SPSS Statistics 23 package. While GIP species diversity was defined as the total number of parasite species detected during the study, GIP prevalence was defined as the proportion of parasite infected bats to the total number examined. Species richness meant the number of different GIPs in bats per site, and evenness meant the relative abundance of GIPs detected in bats per site, and these were calculated using the Shannon-Wiener diversity index (Table 2). The Pearson chi-square test was used to assess any statistical difference in the overall GIP prevalence rate between micro- and mega-bats in Burondo sub-county and Ntandi town council. Mann-Whitney *U* test was used to assess any statistical difference in the overall GIP prevalence rate between micro- and mega-bats during the study. Confidence intervals (95%) and P < 0.05 were set for significance.

## Results

Bat faecal samples (n=484): 242 from Burondo sub-county and 242 from Ntandi town council (Table 1) were tested for parasite eggs, cysts, and/or oocytes using standard floatation and sedimentation diagnostic techniques. Thirteen parasite species, including protozoa (n=3), trematode (n=1), cestode (n=1), and nematodes (n=8) were detected (Table 1). Of the 484 bat faecal samples examined, 224 (46%) tested positive for more than one parasite species. Among the detected parasite species, seven were shared across the four bat groups (Table 1). The most prevalent parasites were *Entamoeba coli* (57%), Hookworm (33%), *Strongyloides* sp. (33%), and *E*. *histolytica* (32%), and the least prevalent were the two Unidentified nematodes (1%) (Table 1).

**Table 1:** Prevalence (%) of gastrointestinal parasites diagnosed in bat guano in Burondo sub-county and Ntandi town council, Bundibugyo district western Uganda.

| Parasite | Burondo sub-county |  | Ntandi town council |  | Overall prevalence (%) | Chi-square test |  |
| --- | --- | --- | --- | --- | --- | --- | --- |
|  | Microbats | Megabats | Microbats | Megabats |  | X<sup>2</sup> | P value |
|  | (n=121)<br>n (%) | (n= 121)<br>n (%) | (n= 121)<br>n (%) | (n= 121)<br>n (%) |  |  |  |
| **Protozoa** |  |  |  |  |  |  |  |
| *Entamoeba coli* | 81 (67.8) | 67 (55.4) | 72 (59.5) | 55 (45.5) | 57 | 11.88 | 0.008 |
| *E. histolytica* | 37 (30.6) | 43 (35.5) | 50 (41.3) | 26 (21.5) | 32 | 11.43 | 0.01 |
| *Eimeria* sp. | 19 (15.7) | 13 (10.7) | 20 (16.5) | 11 (9.1) | 13 | 4.29 | 0.232 |
| **Trematoda** |  |  |  |  |  |  |  |
| *Fasciola* sp. | 10 (8.3) | 0 (0.0) | 3 (2.5) | 0 (0.0) | 3 | 21.11 | *** |
| **Cestoda** |  |  |  |  |  |  |  |
| *Hymenolepis* sp. | 21 (17.4) | 0 (0.0) | 5 (4.1) | 0 (0.0) | 5 | 48.29 | *** |
| **Nematoda** |  |  |  |  |  |  |  |
| Hookworm | 59 (48.8) | 19 (15.7) | 72 (59.5) | 9 (7.4) | 33 | 104.41 | *** |
| *Trichuris* sp. | 37 (30.6) | 9 (7.4) | 33 (27.3) | 11 (9.1) | 19 | 34.67 | *** |
| *Ascaris* sp. | 43 (35.5) | 14 (11.6) | 36 (29.8) | 5 (4.1) | 20 | 49.39 | *** |
| *Strongyloides* sp. | 68 (56.2) | 22 (18.2) | 53 (43.8) | 19 (15.7) | 33 | 63.72 | *** |
| *Angiostrongylus* sp. | 11 (9.1) | 2 (1.7) | 7 (5.8) | 0 (0.0) | 4 | 15.44 | 0.001 |
| *Chitwoodspirura* sp. | 10 (8.3) | 1 (0.9) | 2 (1.7) | 0 (0.0) | 3 | 19.84 | *** |
| Unidentified nematode 1 | 2 (1.7) | 0 (0.0) | 1 (0.9) | 0 (0.0) | 1 | 3.69 | 0.297 |
| Unidentified nematode 2 | 0 (0.0) | 0 (0.0) | 4 (3.3) | 0 (0.0) | 1 | 12.1 | 0.007 |

Key: n=number of bat faecal samples examined, sp. = species, %= percentage prevalence, $\chi^2$ = Chi-square value, P-value = Chi-square P-value, \*\*\* = P-value less than 0.001

While the parasite species diversity ranged between 1.67 ≤ H’ ≤ 2.20, parasite species richness ranged from 53.8% to 100%, and parasite species evenness was 0.65 ≤ J’ ≤ 0.86 (Table 2). The parasite species diversity and evenness lie within the normal standard range of 1.5 ≤ H’ ≤ 3.5 and 0 ≤ J’ ≤ 1, respectively, indicating uniformity in the four studied bat groups [42, 43]. Statistically, a significant difference, *X*^2^(3, N = 484) = 213.2, *P* < 0.05, was seen in the overall parasite prevalence across the four studied bat groups. The Mann-Whitney *U* test revealed that seasonality did not have any influence on parasite prevalence (*P >* 0.05) and that parasite prevalence was significantly higher in microbats compared to megabats, *P* < 0.05, and significantly higher in Burondo sub-county compared to Ntandi town council, *P* =0.002 (*P* < 0.05).

**Figure 3:**
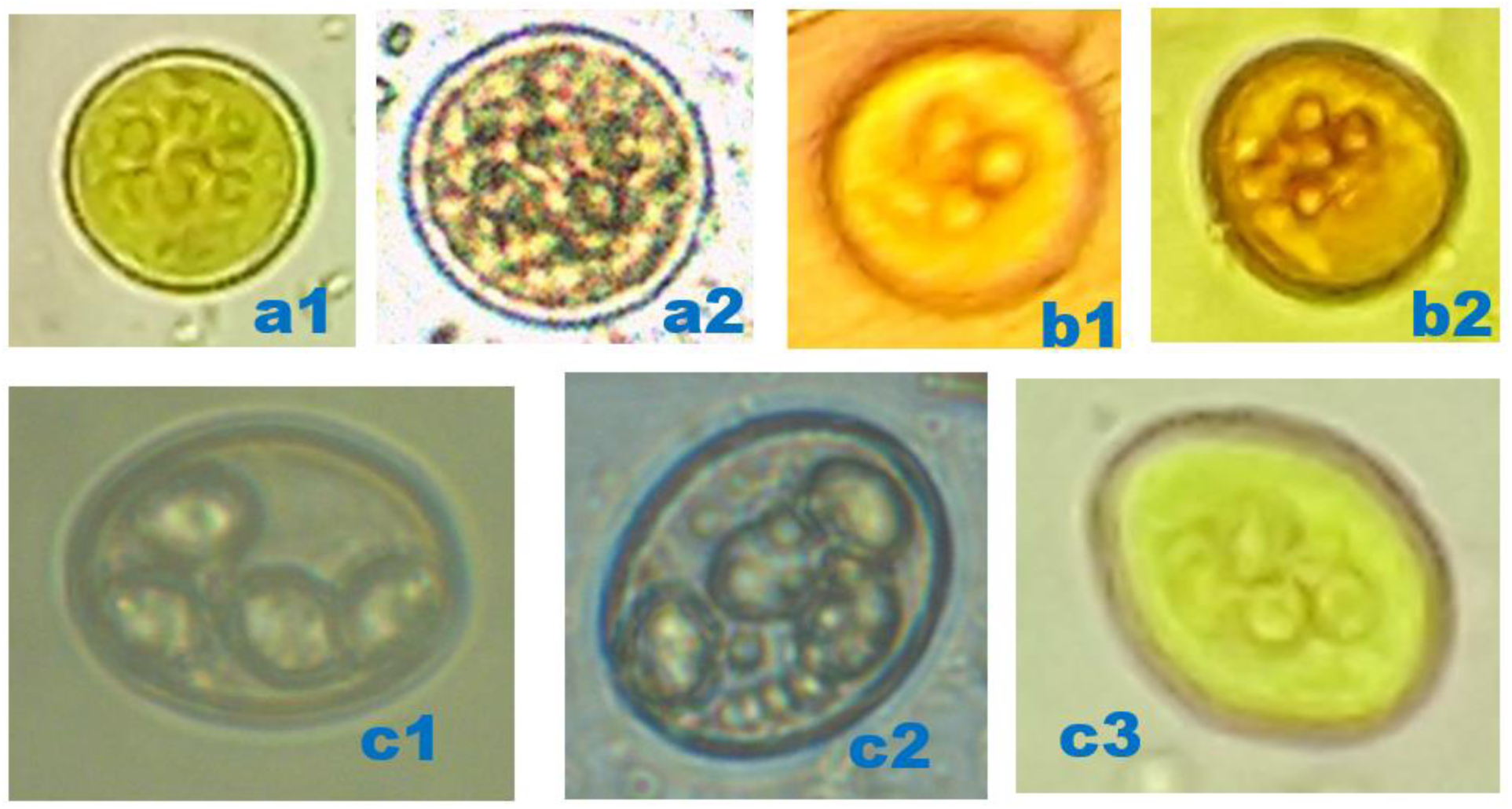
Photomicrographs of protozoan parasites detected in bats. a1) *Entamoeba coli* cyst (10 x 10 µm), 400x, after floatation technique, in microbat. a2) *E*. *coli* cyst (12 x 12 µm), 400x, after floatation technique, in megabat. b1) *E*. *histolytica* cyst (11 x11µm) with four nuclei, 400x, after floatation technique, in microbat. b2) *E*. *histolytica* cyst (10 x 10 µm) with four nuclei, 400x, after floatation technique, in megabat. c1) *Eimeria* sp. oocyst (14 x 14 µm) with four sporocysts, 400x, after floatation technique, in microbat. c2) *Eimeria* sp. oocyst (14 x 14 µm) with four sporocysts, 400x, after floatation technique, in megabat. c3) *Eimeria* sp. oocyst (14 x 14 µm) with four sporocysts, 400x, after sedimentation technique, in microbat. *Original micrographs by James Robert Ochieng*.

**Figure 4:**
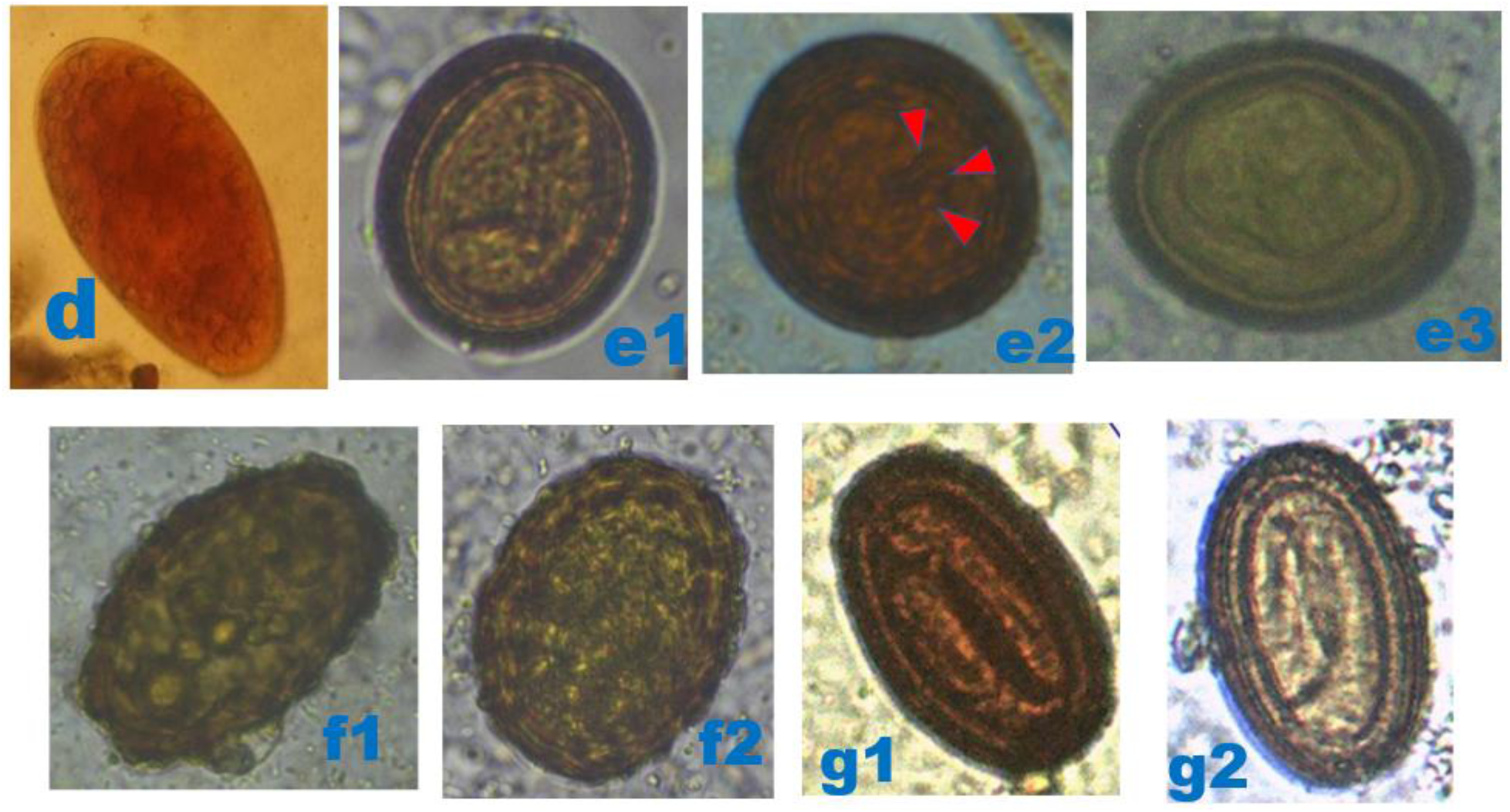
Photomicrographs of helminthic parasites detected in bats. d) Ellipsoidal *Fasciola* sp. egg (95 x50 µm), 400x, after sedimentation technique, in microbat. e1 and e2) large round *Hymenolepis* sp. egg (52 x 52 µm), 400x, after floatation technique, in microbat, the red arrowheads show the 3 small paired hooks e3) Oval *Hymenolepis* sp. egg (45 x 35 µm), 400x, after floatation technique, in microbat. f1) Unfertilized *Ascaris* sp. egg (75 x 40 µm), 400x, after floatation technique, in microbat. f2) Fertilized *Ascaris* sp. egg (65 x 60 µm) with a thick shell having an external mamillated layer/ rippled surface, 400x, after floatation technique, in megabat. g1 and g2) *Chitwoodspirura* sp. eggs (70 x 45 µm), 400x, after floatation technique, in microbat and megabat, respectively. *Original micrographs by James Robert Ochieng*.

**Figure 5:**
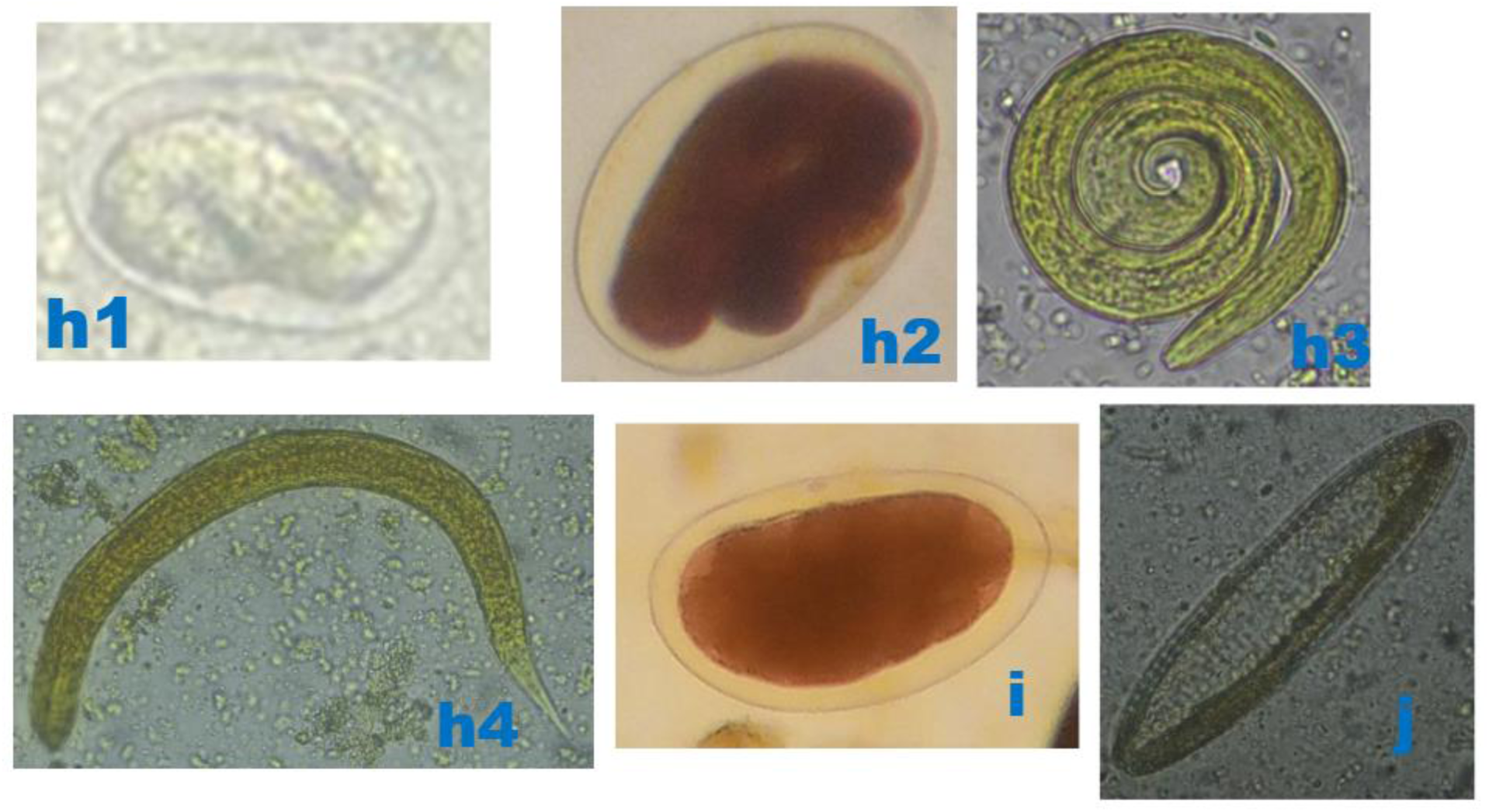
Photomicrographs of helminthic parasites detected in bats. h1) Thin walled larvated/embryonated ellipsoidal egg of *Strongyloides* sp. (80 x 65 µm), 400x, after floatation technique, in microbat. h2) Thin walled larvated ellipsoidal egg of *Strongyloides* sp. (86 x 66 µm), 400x, after floatation technique, in megabat. h3 and h4) Coiled and stretched larvae of *Strongyloides* sp., 400x, after floatation technique, in microbat and megabat, respectively. i) Unidentified nematode 1 egg (68 x 45 µm), 400x, after floatation technique, in microbat. j) Unidentified nematode 2 egg (78 x 30 µm), 400x, after sedimentation technique, in microbat. *Original micrographs by James Robert Ochieng*.

**Figure 6:**
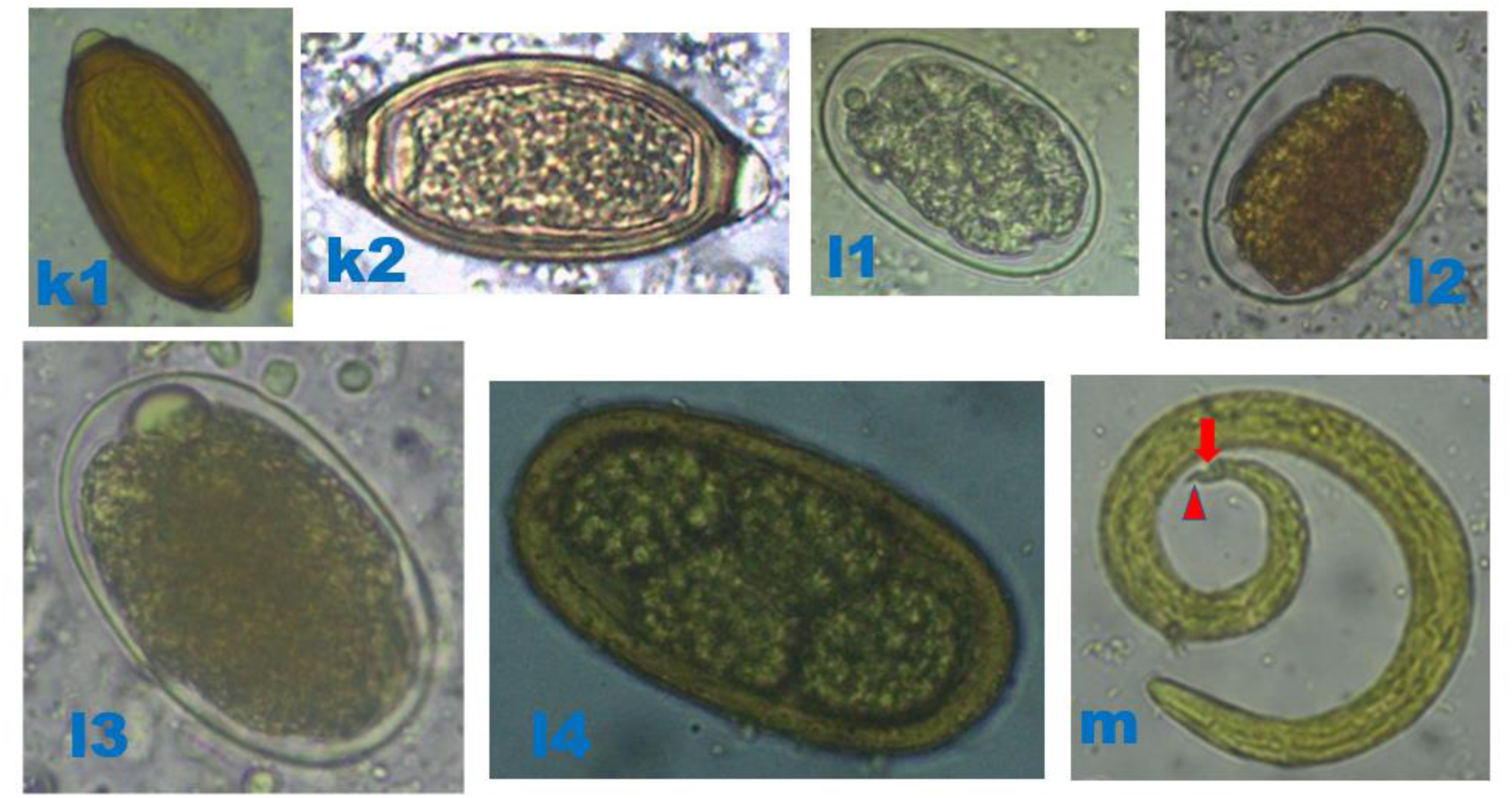
Photomicrographs of helminthic parasites detected in bats. k1) Brown barrel-shaped *Trichuris* sp. egg (50 x 25 µm), 400x, after sedimentation technique, in microbat. k2) Barrel-shaped *Trichuris* sp. egg (54 x 30 µm), 400x, after floatation technique, in megabat. Both k1 and k2 possess a thick shell and a pair of polar plugs (bipolar protuberances) at each end. l1 and l2) Thin smooth oval-shelled hookworm eggs (55 x 35 µm), 400x, after floatation technique, in microbats and megabats, respectively. l3) Oval thin smooth-shelled hookworm egg (75 x 45 µm), 400x, after floatation technique, in megabats. i4) Oval hookworm egg (77 x 40 µm) with four grouped cells “morula”, 400x, after sedimentation technique, in megabats. m) *Angiostrongylus* sp. larvae with a dorsal spine and notch (red arrow), and tail terminates in wave-shaped kink (red arrowhead), 400x, after sedimentation technique, in microbats. *Original micrographs by James Robert Ochieng*.

**Table 2:**
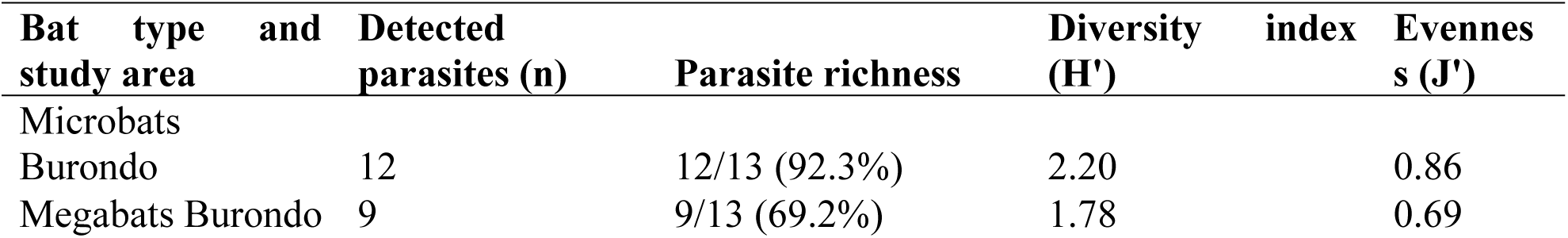
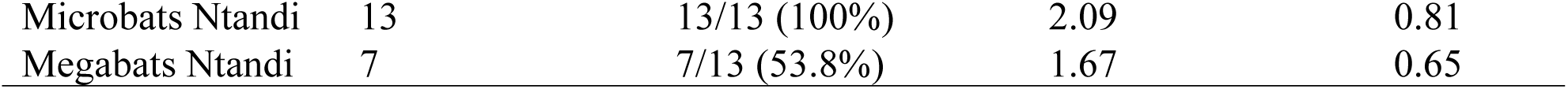
Bat Gastrointestinal parasite richness, diversity, and evenness in micro- and mega-bats in Burondo sub-county and Ntandi town council, Bundibugyo district western Uganda.

## Discussion

From the parasitological point of view, bats are of special interest for several reasons. Bats are susceptible to many infections of animals and humans, thus are important vectors and/ or reservoirs of pathogens of public health importance [6, 8, 37]. This study determined the diversity and prevalence of bat GIPs in human communities contiguous to SNP, Bundibugyo district. To our knowledge, this is the first extensive research to assess bat GIPs in communities contiguous to protected areas of SNP and Uganda. Of the detected parasites, 77% (n = 10) are zoonotically important in the aspect of public health but also important concerning bat /wildlife conservation efforts [6, 8]. To date, there is still little information on the diversity and prevalence of GIPs on several mammalian species of wild fauna despite the currently recognized increase in human-wildlife interaction in the study area [19]. The present study findings are critical in understanding the prevalent parasites that affect the bats’ health in the study area and the possible zoonotic agents they harbour.

The overall bat GIP prevalence was 46%. This is low compared to 63.6%, 76%, 76.78%, 80%, and 96.29% detected in Arkansas and North Carolina [32], England [44], Nigeria [27], Nepal [15], and Brazil [45], respectively. However, the detected bat GIP prevalence in the present study is higher compared to other previous related findings, namely, [26] reported 43.9 % GIPs in bats in Egypt, [46] reported 14 % GIPs in bats in Costa Rica, and [47] reported zero (0 %) prevalence in bats in Ghana.

The recognized differences in GIP prevalence in the current study to previous ones could mostly be attributed to a combination of factors, including but not limited to varying climatic conditions and temperatures and environmental factors like sanitation that influence the survival, development, and spread of parasites [6, 37]. For example, GIPs survive better in the tropical and sub-tropical regions conducive for the development and transmission of parasitic larvae than in the temperate climates [6]. Other factors may include GIP species and their spread mechanisms; guano sample size: larger sample size may have higher chances of more GIP diversity and prevalence; and bat types and their feeding characters: insectivorous bats encounter and eat many GIP reservoirs unlike the megabats [8, 37].

Studies have also shown that bats’ GIP diversity and prevalence are correlated with their eating habits [26, 48]. Previous related studies by [48] and [26] have shown that insectivorous bats acquire parasites by eating GIP reservoirs, including insects (like cockroaches, dung beetles), amphibians, rodents, snails, and raw fish [6, 8]. This is the opposite of megabats/ fruit eaters [26]. Furthermore, only a few or no cases of trematode have been detected in fruit bats, including *Rousettus aegyptiacus*, unlike in insectivorous bats [26, 49]. Indeed, the present finding is in line with these previous related studies, where parasite richness was higher in insectivorous bats (92.3-100%) than in megabats (53.8-69.2%). Also, the overall GIP prevalence was higher in insectivorous (65 %) than megabats (28 %), and only insectivorous bats tested positive for trematode eggs, as in previous related studies by [49] and [26].

In the present study, the highly and moderately prevalent parasites, namely, *E*. *coli*, *E. histolytica, Eimeria* sp., hookworms, *Strongyloides* sp., *Ascaris* sp., and *Trichuris* sp., are known to have several reservoir hosts [6, 21, 37]. Some also have horizontal transmission, for example, *E*. *coli*, *E. histolytica, Eimeria* sp., and *Trichuris* sp. [21]. This is opposite for the low prevalent GIPs like *Fasciola* sp. and *Chitwoodspirura* sp. with indirect life cycles and fewer intermediate hosts [6, 21].

GIP horizontal transmission in bats occurs through the direct ingestion of contaminated food and/ or water [8, 37]. Insectivorous bats acquire *Fasciola* sp. by ingestion of infected snails, the intermediate host during feeding [6]. In a similar case, insectivorous bats acquire *Chitwoodspirura* sp. through ingesting bat flies (families: Nycteribiidae and Streblidae), the obligate ectoparasites of bats, but also intermediate hosts for *Chitwoodspirura* sp. [6]. The recognized *Hymenolepis* sp. parasites in the insectivorous bats could be attributed to their eating of rodents, known to be both reservoir and definitive host, but also eating of arthropods, like beetles, the reservoir host [21, 37]. In addition, *Hymenolepis* sp. also has direct transmission and autoinfection competency [21]. *Angiostrongylus* sp. (family Metastrongylidae) is a parasitic nematode with two species, *A*. *cantonensis* (rat lungworm) and *A*. *costaricensis*, causing human infections. The former causes eosinophilic meningitis and later leads to abdominal angiostrongyliasis in humans [50] and could be the same in bats. Insectivorous bats acquire *Angiostrongylus* sp. by eating infected snails or slugs, the intermediate hosts [6, 50], though it is not clear how megabats acquire this parasite as in the present study.

The two Unidentified nematodes were only detected in microbats. The Unidentified nematode 1, marked as micrograph “i”, appeared in the shape of *Enterobius* species parasite, a known GIP with a zoonotic potential [6]. Also, the Unidentified nematode 2, marked as micrograph “j”, appears in the shape of *Oxyurid* sp. egg as it was the case in insectivorous bats in Southcentral Nepal by [15], but more interventions are still needed to classify it satisfactorily. Nonetheless, *Oxyurid* species are known GIPs of rodents, and insectivorous bats can acquire infections by eating infected rodents [6].

The detected parasites in this study can negatively impact bats’ health, reproductive success, and survival, potentially leading to malnutrition and increased disease susceptibility, and death [51–53]. The detected bat-borne zoonotic parasites pose a risk to human health [7, 21, 54], highlighting the importance of understanding the interactions between bats, parasites, and humans in the study area and beyond.

### Limitations and recommendations

Though the risk factor data designed with questionnaires capturing community awareness, knowledge, and attitudes on bat mediated zoonosis in the study area was captured in another related study report [19, 55], the current study has limitations that might have affected the results. The first limitation relates to the taxonomy of the studied bats; we did not classify the bats to species level, though we were able to categorize them as micro- and mega-bats. Secondly, we did not use molecular techniques during parasite diagnosis, so we could not classify most of the parasites to species level.

Based on the currently increasing changes in habitat, forest fragmentation, and urbanization, screening wildlife populations for zoonotic parasites is crucial for public health safety. Nevertheless, studying parasites associated with wildlife offers a significantly different scenario to that of humans and/or domestic animals and contributes to wildlife conservation.

Due to the current findings, future studies could benefit from classifying the bats and GIPs to species level using molecular techniques. This will shed more light on differentiating the bats and the GIP species. Such data is crucial in public health and biodiversity conservation in Uganda and beyond.

### Conclusions

This study confirms the circulation of bat mediated zoonotic parasites in human communities contiguous to SNP, which poses public health risks. This study, being the first of its kind in Uganda, addresses the knowledge gap in bat GIPs. Therefore, this data highlighted the critical role of bats as sentinel species for effective GIP surveillance in bat hotspot areas. This enhances the need for early detection, control, and prevention of parasite zoonosis in human communities, especially in contiguity and/ or proximity to the protected areas known to possess many reservoir hosts.

## Data Availability

All data produced in the present study are available upon reasonable request to the authors

## Acknowledgments

We would like to thank the District Veterinary Officers of Bundibugyo district, the Village Health Teams, and the community members in the studied areas for their significant voluntary assistance during the data collection processes.

## Author contributions

JRO and CDK conceived and designed this study, carried out field data collection and guano sampling. JRO conducted laboratory and statistical analyses. JRO, CDK, and JJK wrote the first draft of the manuscript. All authors have read and approved the manuscript for submission for publication.

## Data availability

The data supporting this study’s findings are available from the corresponding author upon request.

## Declarations

### Ethical approval

The study experimental plan was conducted following the Ministry of Agriculture, Animal Industry and Fisheries (MAAIF), Uganda, Uganda Wildlife Authority (UWA), and Ministry of Health (MOH), Uganda guideline and requirement for wildlife conservation and human community in Uganda. Therefore, this was a public health control program and did not require ethics committee approval or written consent from communities in the study area.

### Consent to publish

Not applicable.

